# Covid-19 mortality rates in Northamptonshire UK: initial sub-regional comparisons and provisional SEIR model of disease spread

**DOI:** 10.1101/2020.07.30.20165399

**Authors:** Nick Petford, Jackie Campbell

## Abstract

**Objectives:** We analysed mortality rates in a nonmetropolitan UK subregion (Northamptonshire) to understand SARSCoV2 disease fatalities at sub 1,000,000 population levels. A numerical (SEIR) model was then developed to predict the spread of Covid19 in Northamptonshire.

**Methods:** A combined approach using statistically-weighted data to fit the start of the epidemic to the mortality record. Parameter estimates were then derived for the transmission rate and basic reproduction number.

**Results:** Age standardised mortality rates are highest in Northampton (urban) and lowest in semi-rural districts. Northamptonshire has a statistically higher Covid-19 mortality rate than for the East Midlands and England as a whole. Model outputs suggest the number of infected individuals exceed official estimates, meaning less than 40% of the population may require immunisation.

**Conclusions:** Combining published (sub-regional) mortality rate data with deterministic models on disease spread has the potential to help public health practitioners develop bespoke mitigations, guided by local population demographics.

## Introduction

Since the global outbreak of SARS-CoV-2 in December 2019, the UK has been one of the hardest hit nations in reported mortality rates, with deaths by 1 million of the population second only to Belgium. Lockdown in the UK started on March 24 2020, several weeks later than elsewhere in mainland Europe. The UK response has been criticised by some for lagging, despite early mortality data from Hubei, China, South Korea and Italy, where sufficient data have been gathered to model the initial spread of the virus^1,2,3^.

Despite this, it is important not to rely entirely on data aggregated at national level to make local public health interventions, once the virus has taken a steady hold. Given the duration since the first recorded deaths in the UK, the opportunity now presents itself to look more closely at regional and sub regional trends that impact on location-specific public health mitigations^4^. The aim of this research is to analyse in detail mortality rates due to Covid-19 in a non-metropolitan UK subregion (Northamptonshire) to understand SARS-CoV-2 disease fatalities at granular (< 1 million) population level. Northamptonshire was chosen as case study because while largely a rural county, it has significant centres of urban population with mixed ethnicity, typical of English counties more generally. The detailed comparisons made on mortality rates (and place of death) are a useful catalogue in themselves. In addition, they provide robust input for deterministic models seeking to understand how the virus is spread.^2^

This paper comprises two parts: 1. An analysis of publicly available mortality data in Northamptonshire benchmarked nationally and regionally, and 2) a SEIR mathematical model that applies these data to better understand viral spread locally. Our goal is to combine observations with mathematical models to help prepare for better long-term regional planning in event of a second (or move) waves of Covid-19, should it occur.

## Methods

Data on age standardised mortality rates involving Covid-19 form the period March to June 2020, published on July 24 by the ONS for each district and local authority in Northamptonshire, were analysed^5^. The rates between geographical areas were different (p<0.05) if there was no overlap in the 95% confidence intervals for the comparison areas. All data used in this study are publicly available and licenced for use under ONS Open Access rules. The SEIR model was run using COMSOL v. 5.5 Multiphysics (Finite Element) software, calibrated against weekly ONS death rates by occurrence in Northamptonshire.

## Regional Mortality Rates

### East Midlands

The East Midlands is one of nine official regions of England at the first level of NUTS (The Nomenclature of Territorial Units for Statistics) for statistical purposes. It consists of six English counties, including Northamptonshire, with a combined population of 4.811 million^6^. The largest city by population is Leicester, site of the first sub-national lockdown in the UK. Northampton is the fourth largest conurbation in the region. A time-series comparison of national to sub-regional mortality rates, is shown in Fig. 1.

**Fig. 1.**
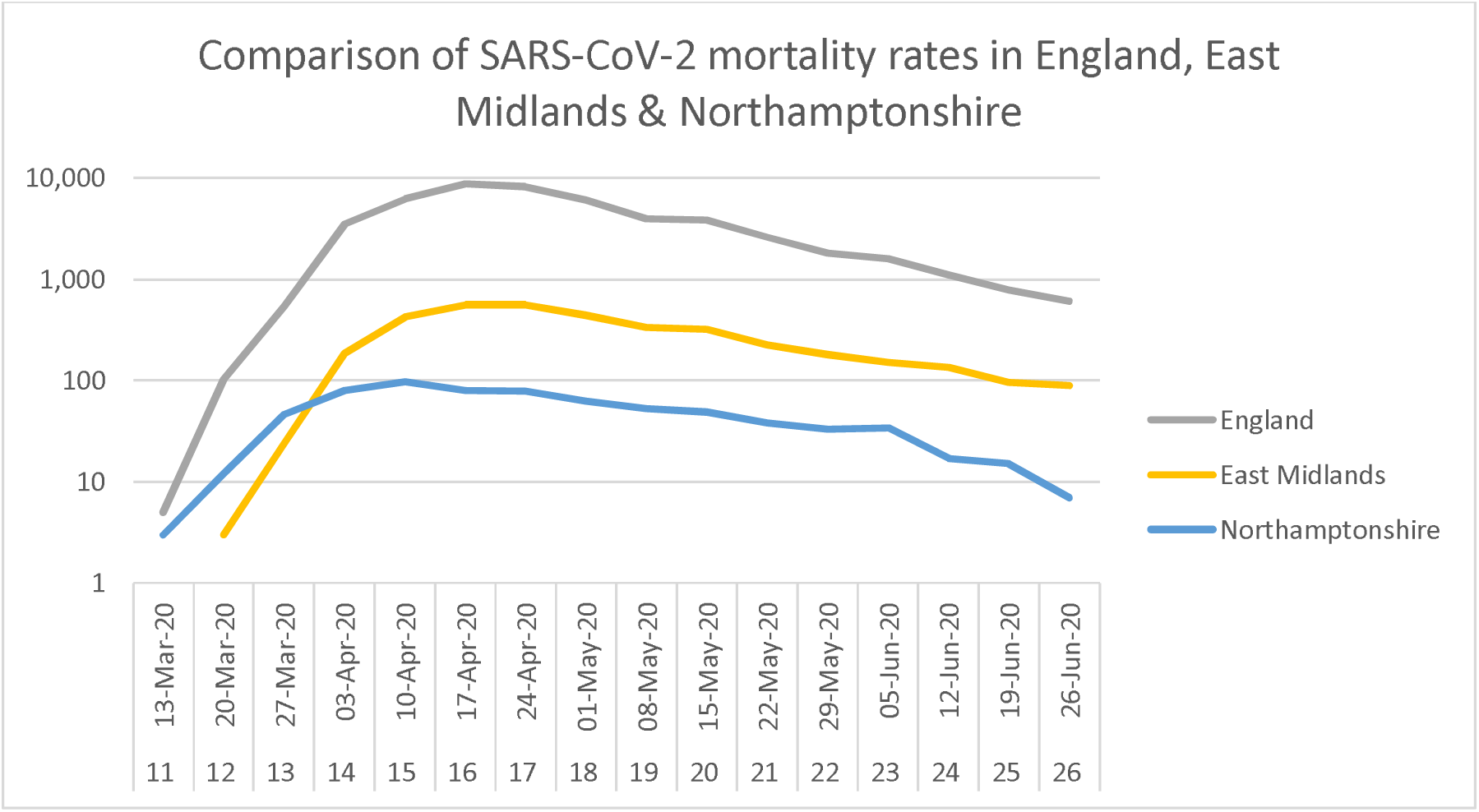
Log_10_ of registered mortality rates in England, East Midlands and Northamptonshire weeks 11-26, 2020^5^ showing early characteristic linear (log exponential) increase^7^.

### Northamptonshire

Northamptonshire (population 753,278^6^, density 316 km squared), is the southernmost county in the East Midlands region, covering an area of 2,364 square kilometres. The major urban centre is Northampton (population, 224,610^6^). The 2011 census showed the county split evenly between males (49.5%) and females (50.5%), with 18.1% of the population aged 65 years and older. Nearly 90% of the population are white (British or other). In terms of governance, Northamptonshire is currently divided into seven boroughs and local district councils following a two-tier structure of local government. In March 2018, new structural changes were proposed that see the existing boroughs and district councils replaced by two unitary authorities of West and South Northamptonshire.

## Analysis

### Comparison of age-standardised Covid-19 death rates between Northamptonshire areas

Registered deaths due to Covid-19 in Northamptonshire over the current period of study are 715, comprising approximately 25% of total registered deaths over the same period. The data have been broken down by region and district to allow comparisons. The first recorded Covid-19 death was on March 17 (t = 0, week 11), peaking individually at total 98 registered deaths in week 15 (April 13-19). Regarding place of death, the majority (c. 72%) occurred in hospital. When disaggregated into the seven comprising districts it becomes clear that Northampton is the dominant contributor to the overall curve profile with the highest Covid-19 mortality rate (Table 1). This is statistically different, at the 5% level, from all other local authority areas except for Corby. South Northamptonshire has the lowest Covid-19 mortality rate, although only the differences with that of Northampton and Kettering are statistically significant. Compared regionally, Northamptonshire has a statistically higher Covid-19 mortality rate than for the East Midlands as a whole. Both the East Midlands region and Northamptonshire have statistically significantly higher rates than for the whole of England (Fig. 2 and Table 1).

**Table 1.** Covid-19 mortality rates by area for local authorities in Northamptonshire with regional comparators and new Unitary Authority areas. Data for all sexes for the combined months March-June 2020^5^. Age standardised to European Standard Population (ESP) (2013). Population estimates are for mid-2019^6^.

| Area | No. deaths | Population estimate | Age-standardised rate (/10,000) | Lower 95% CI | Upper 95% CI |
| --- | --- | --- | --- | --- | --- |
| Corby | 50 | 72,218 | 101.4 | 74.9 | 134.1 |
| Daventry | 75 | 85,950 | 88.4 | 69.4 | 110.9 |
| E Northamptonshire | 96 | 94,527 | 96 | 77.7 | 117.3 |
| Kettering | 99 | 101,776 | 104.7 | 85.1 | 127.5 |
| Northampton | 269 | 224,610 | 148 | 130.2 | 165.7 |
| S Northamptonshire | 56 | 94,490 | 60.7 | 45.8 | 78.9 |
| Wellingborough | 66 | 79,707 | 87.8 | 67.9 | 111.8 |
| Northamptonshire | 711 | 753,278 | 105 | 97.2 | 112.7 |
| East Midlands | 3812 | 4,835,928 | 79.9 | 77.3 | 82.4 |
| England | 48040 | 56,286,961 | 88.7 | 87.9 | 89.5 |
| Unitary Authority North | 311 | 348,228 | 97.5 | 86.6 | 108.3 |
| Unitary Authority West | 400 | 405,050 | 99.0 | 89.3 | 108.7 |

**Fig. 2.**
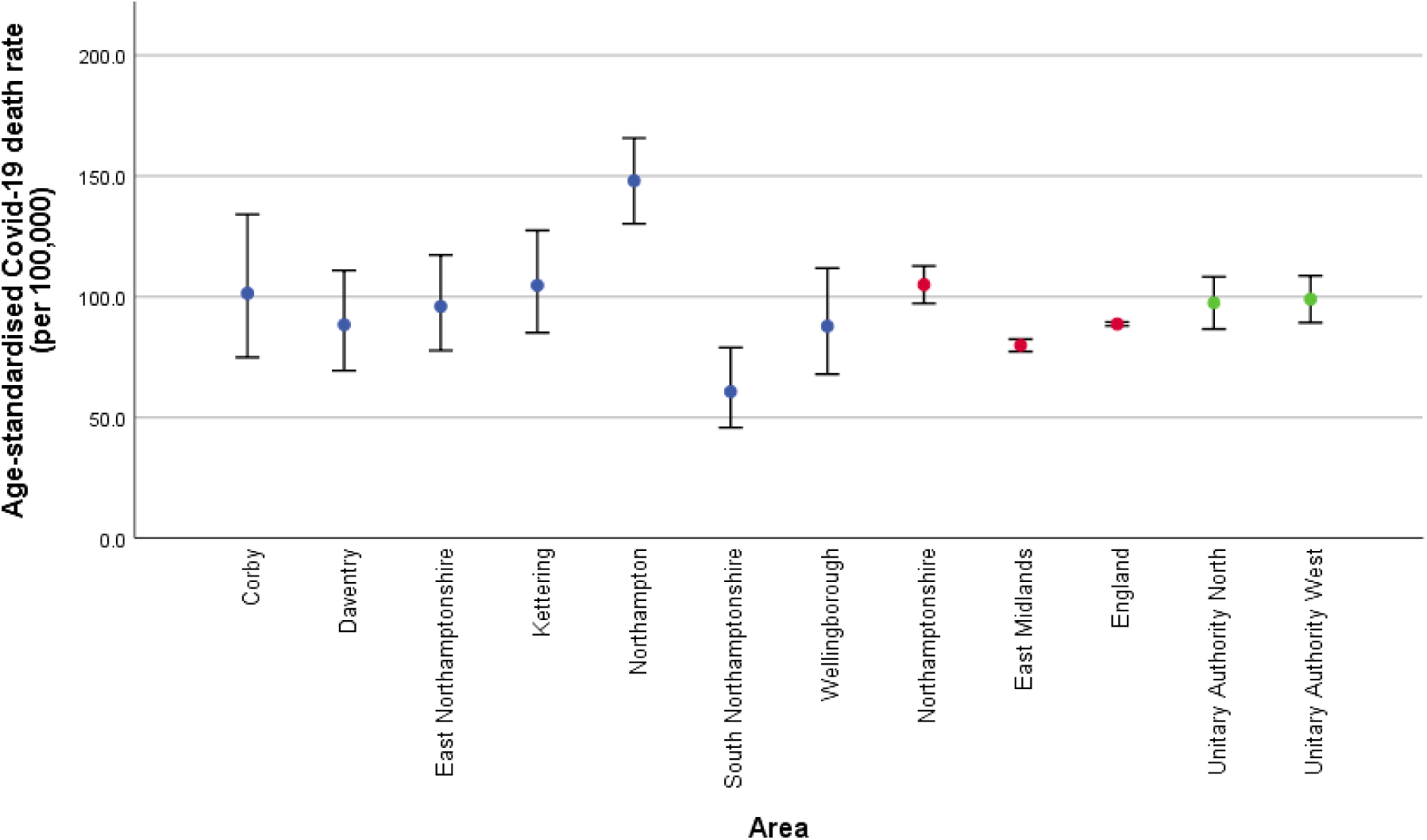
Age-standardised death rates (per 100,000) from Covid-19 for all sexes for the combined months of March-June 30 2020.^5^ Blue circles indicate the rates for the current Northamptonshire local authorities, red circles denote regional comparators and green circles are the rates for the new Unitary Authority areas. Age standardised to European Standard Population (ESP) (2013).

The new political structure, due to take effect in April 2021, has consequences for the future management and resourcing of public health in its ambition to reduce health inequalities. However, if the new Unitary Authorities were in place, then there would be no statistically significant difference in the Covid-19 mortality rates between them. This is because Northampton, with the highest rate in the county, and South Northamptonshire, with the lowest, are both in the West Northamptonshire Unitary Authority.

### Provisional SEIR model for Northamptonshire

One of the simplest mathematical techniques used to predict disease transmission is the SEIR (or SIR) model^7,8,9^ which divides a population N at t = 0 into four ‘compartments’ of susceptible S(t), exposed (meaning infected but not yet infectious) E(t), Infectious, I(t) and recovered R(t). Individuals move between compartments in the model at a rate determined by four interlinked ordinary differential equations such that S(t) + E(t) + I(t) + R(t) = N. SEIR models are deterministic and average the infectiousness across a susceptible population. They are thus different from stochastic models where infection is modelled via discrete interactions between individuals^10,11^. Key variables in both include the transmission rate by infectious individuals (*β*), the average number of days a person is infectious (n), and the recovery rate γ (1/n). They are related via: *β* = R_0_/n, where R_0_ is the basic reproduction number. Any mitigation strategy must aim to reduce the reproduction number, for example by decreasing the transmission rate or the time infections individuals are isolated^12^.

Current uncertainties in the modelling relate explicitly to the details of disease transmission. Covid-19 is transmitted from symptomatic (infected) people through respiratory droplets or contact with contaminated materials^13^. The infectious period (n) appears maximised in the first three days after infection. The incubation period for COVID-19, which is the time between exposure and symptom onset is 5-6 days on average but can be up to 14 days^14^. During this time (pre-symptomatic) period, an unknown fraction of infected individuals may be contagious, meaning transmission can occur before symptom onset. The model assumes homogenous mixing within the population and does not account for asymptomatic transmission. Imported cases are also excluded, although provision exists to add this variable in future modelling.

Two scenarios are presented, one where the virus is left to run its course after the first registered deaths at t = 0 for a period of 120 days. The second model (with restrictions), follows the UK Government response with lock down mitigations imposed on March 24, seven days later. It mimics the effects of social distancing from t = 7 onwards by reducing the transmission rate and basic reproduction number relative to the ‘no restrictions’ case by 40% (Table 2). Although the results are provisional and subject to revision as new data emerge (meaning the parameterised fit to the mortality data may change), they nonetheless provide information about disease veracity and spread useful for guiding public health mitigation at sub-regional level^4^ (see discussion).

**Fig. 3.**
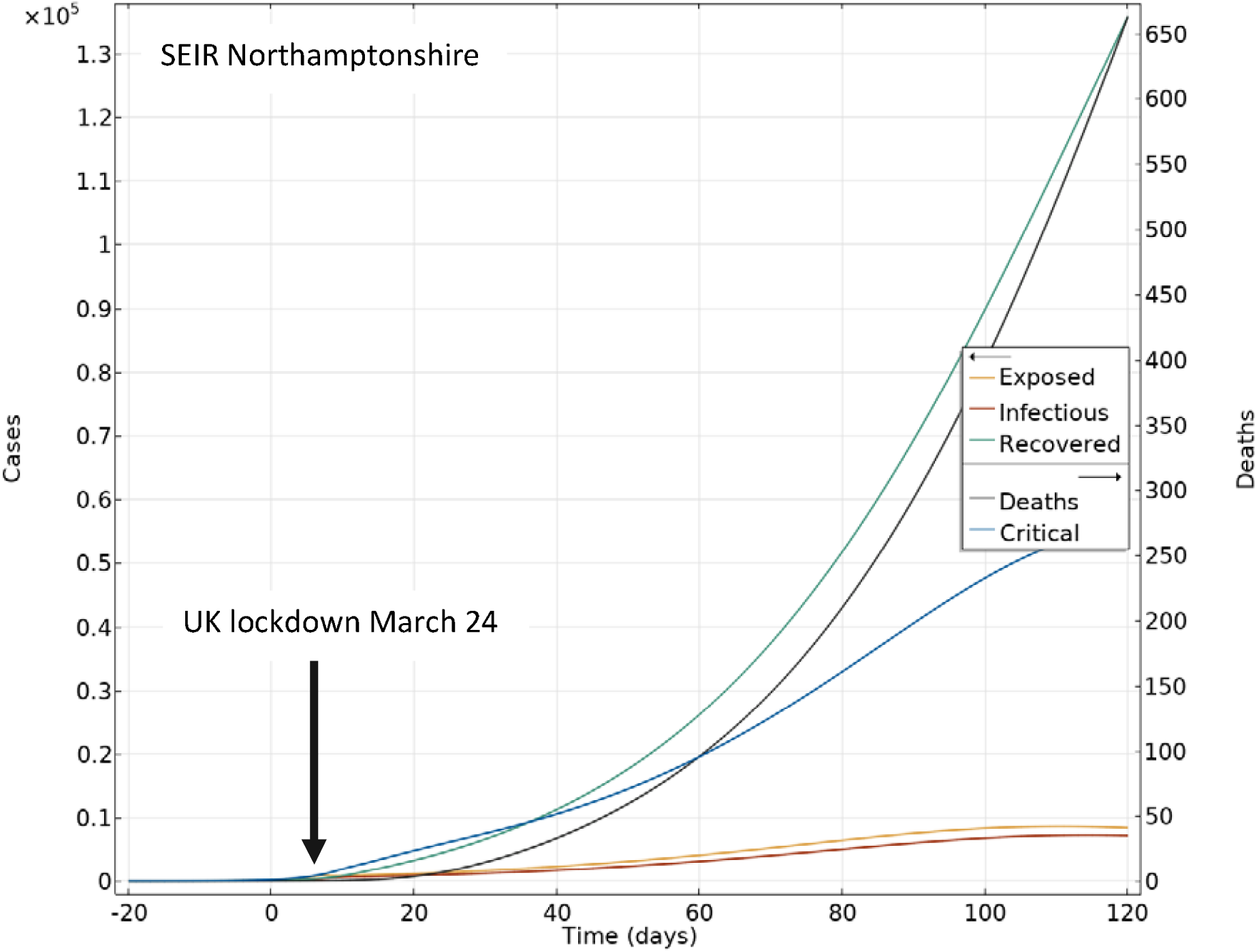
SEIR ‘with reductions’ model for Northamptonshire calibrated against mortality rate (Fig 2), showing numbers of cases and deaths, along with exposed, infectious and recovered. Reductions start (March 24) at day 7 from t = 0. Simulation time 120 days. Note the slight ongoing rise in infections (R0 > 1, Table 2).

**Table 2.** Summary of input data^1^ and results from SEIR model for Northamptonshire assuming import cases = 0, Initial terminally ill state mean residence time = 18 days^14^.

| Model inputs | Symbol/expression | SEIR 'no reductions' | SEIR 'with reductions' |
| --- | --- | --- | --- |
| Population | N | 7.533E+05 | 7.533E+05 |
| Infectious at t = 0 | I(t = 0) | 3 | 3 |
| Transmission rate (day) | ($\beta$) | 0.98 | 0.59 |
| End of simulation time (days) | t | 120 | 120 |
| Days infectious | n | 3 | 3 |
| Recovery rate from infection | $\gamma=1/n$ | 0.33 | 0.33 |
| Total recovered | R(t) | 7.01E+05 | 1.36E+05 |
| Total deaths | D | 4585 | 663 |
| Basic reproduction number | R<sub>0</sub> | 2.98 | 1.21 |
| Critical vaccination threshold %* | $\rho_c$ | 67 | 36 |

## Results

Results from the SEIR models are summarised in Table 2. The basic reproduction number (2.9) in the ‘no-reductions’ case lies within the reported range for Covid-19^15^, consistent with early exponential spread of the virus^7^. The simulations highlight the positive impact of reductions on the mortality rate. Allowed to spread uninhibited, total predicted deaths exceed 4500 against an actual registered of 715, c. six times higher than currently recorded. In contrast, the ‘with reductions’ scenario predicts 633 deaths over the same period. The potential number of infected individuals is reflected in the number of recovered (Rt) cases. The ‘with reductions’ model implies 135,800 discrete exposures to the virus over the simulation period, equating to approximately 18% of the total population. This value is significantly in excess of cases reported for Northamptonshire^16^ and is received with caution. It is however consistent with evidence for significant underreporting of infections during the initial phase of the pandemic. For example, only around 15% of active cases were registered in Wuhan,^17,18^ while SIR modelling by Lourenco et al^19^ suggest the epidemic in the UK started at least a month before the first reported death. If so, that introduces a minimum lag in the Northamptonshire data of t = −78 days, enough in principle to build up a sizeable reservoir of unreported cases.

## Discussion

### Implications for vaccination and herd immunity

We end with a practical example of how SEIR model outputs can be used to help inform public health interventions locally. Predictions of numbers of recovered cases allow estimates of the critical fraction (*ρ*_c_) of the total susceptible population S(t) needed to be vaccinated to ensure herd immunity^8^. This threshold is equal to 1-1/R_0_.

At the onset of a new infection where there are no prior cases, it is reasonable to assume *S(t)* = *ρ*, the proportion that require vaccination. However, the SEIR models reveal a sizeable (hidden) fraction R(t) of recovered cases. These individuals can be excluded, reducing the overall size of the susceptible population by *R(t)/p* = *φ*. Thus, to prevent sustained spread, the revised fraction of the population requiring immunisation must satisfy the inequality:

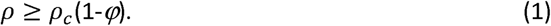

From the SEIR ‘with reductions’ model, *ρ*_c_ is estimated at a provisional 37% of the population (Table 2). Other independent estimates of heard immunity support the idea that Covid-19 critical immunisation thresholds may be substantially less than 50%, especially where some fraction of the population is prevented from transmitting the virus^20^. Should the need arise, statistical analysis (Table 1), suggests Northampton should be treated as a priority target.

We stress again caution is required in when interpreting SEIR model outputs based on data early in the pandemic. As experience from previous epidemics has shown^7,9,12^, initial results may require subsequent revision. Future work will seek to incorporate the geographical and demographic variations identified in the statistical analysis (Table 1) to help refine SEIR models that provide deeper understanding of the dynamics of Covid-19 at sub-regional level.

## Conclusions

Initial results show Northamptonshire mortality rates due to Covid-19 are higher than both regional and national averages. Northampton is the single biggest contributor to mortality rates, with South Northamptonshire the lowest (significance level 0.05). This trend follows current known distributions in health inequalities^16^. A SEIR model, collaborated by fitting to mortality rate, yields predictions about the dynamics of virus spread and local impact of reductions in *β* and R_0_. The introduction of national restrictions (social distancing) may have resulted in up to 4000 fewer deaths in Northamptonshire. Estimates of recovery rates suggest up to 18% of individuals in the county may have been infected, requiring any future immunisation programme to target less than 40% of the Northamptonshire population overall.

## Data Availability

All data used in this study are publicly available, anonymous, and do not require ethical approval. See: ONS Statistical Bulletin. Deaths involving COVID-19 by local area and socioeconomical depravation between March and June 30, 2020. London, July 24, 2020.

https://www.ons.gov.uk/peoplepopulationandcommunity/birthsdeathsandmarriages/deaths/datasets/deathsinvolvingcovid19bylocalareaanddeprivation

## Author Statements

## Ethical Approval

None required. All data are anonymous and publicly available under Open Licence from the UK Office of National Statistics (London)

## Acknowledgements

Lucy Whiteman, Public Health England

## References

1. Verity, R, Okell, LC, Dorigatti, I, Winskill, P, Whittaker, C et al. Estimates of the severity of coronavirus disease 2019: a model-based analysis. The Lancet Infectious Diseases, 2020; 20: 669–677. doi.org/10.1016/S1473-3099(20)30243-7.

2. Kucharski, AJ, Russell, TW, Diamond, C, Liu, Y, Edmunds, J, Funk, S, Eggo, RM. Early dynamics of transmission and control of COVID-19: a mathematical modelling study. The Lancet Infectious Diseases, 2020; 20: 553–558

3. Lee A, Wuhan novel coronavirus (COVID-19): why global control is challenging? Public Health 2020; 179: doi.org/10.1016/j.puhe.2020.02.001.

4. Bray, I, Gibson, A, White, J. Covid-19 mortality: a multivariate ecological analysis in relation to ethnicity, population density, obesity, deprivation and pollution. Public Health, 2020; doi.org/10.1016/j.puhe.2020.06.056.

5. ONS Statistical Bulletin. Deaths involving COVID-19 by local area and socioeconomical depravation between March and June 30, 2020. London, July 24, 2020.

6. ONS, Estimates of the population for the UK, England and Wales, Scotland and Northern Ireland: Mid-2019: April 2020 local authority district codes., ONS, London

7. Chowel G, Sattenspiel, L Bansald, S Viboud, C. Mathematical models to characterize early epidemic growth: A review. Physics of Life Reviews. 2016; 18 :66–97 doi.org/10.1016/j.plrev.2016.07.005.

8. Kermack WO, McKendrick AG. A contribution to the mathematical theory of epidemics. Proc Royal Soc Math Phys Eng Sci. 1927; 115 :700–721.

9. Weiss, H. The SIR model and the Foundations of Public Health. materials matemàtics 2013; 3: 1– 17.

10. Panovska-Griffiths, J. Can mathematical modelling solve the current Covid-19 crisis? BMC Public Health 2020; 20: 551 doi.org/10.1186/s12889-020-08671-z.

11. Ferguson NM, Laydon D, Nedjati-Gilani D, et al. Impact of nonpharmaceutical interventions (NPIs) to reduce COVID-19 mortality and healthcare demand. Preprint assessed 26th March 2020 https://www.imperial.ac.uk/media/imperial-college/medicine/sph/ide/gidafellowships/Imperial-College-COVID19-NPI-modelling-16-03-2020.pdf.

12. Ridenhour, B, Kowalik JM Shay, D., Unraveling R_0_: Considerations for Public Health Applications Am J Public Health. 2014; 104 :e32.–e41.10.2105/AJPH.2013.301704.

13. Zhang, R, Li, Y, Zhang, AL, Wang, Y, Molina MJ. Identifying airborne transmission as the dominant route for the spread of COVID-19. Proc. Nat. Acad. Sciences 2020; 26: 14857–14863; doi:10.1073/pnas.2009637117.

14. Gondauri D, Mikautadze E, Batiashvili M. Research on COVID-19 Virus Spreading Statistics based on the Examples of the Cases from Different Countries. Electron J Gen Med. 2020; 17 :em209. https://doi.org/10.29333/ejgm/7869.

15. Liu, Y, Gayle, AA, Wilder-Smith, A, Rocklöv, J. The reproductive number of COVID-19 is higher compared to SARS coronavirus. J. Travel Medicine, 2020; 7: taaa021, doi:10.1093/jtm/taaa021.

16. Covid-19 Northamptonshire Intelligence Pack, 02/06/2020. Public Health Northamptonshire, Northamptonshire County Council.

17. Li, R, Pei, S, Chen, B, Song, Y et at., Substantial undocumented infection facilitates the rapid dissemination of novel coronavirus (SARS-CoV2). Science, 2020; 368, 489–493 doi:10.1126/scienceabb3221.

18. Wu, J.T., Leung, K., Bushman, M. et al. Estimating clinical severity of COVID-19 from the transmission dynamics in Wuhan, China. Nat Med 2020; 26: 506–510. https://doi.org/10.1038/s41591-020-0822-7.

19. Lourenco, J, Paton, R, Ghafari, M, Kraemer, M, Thompson, C, Simmonds, P, Klenerman, K, Gupta, S. Fundamental principles of epidemic spread highlight the immediate need for large-scale serological surveys to assess the stage of the SARS-CoV-2. 2020; epidemicdoi:https://doi.org/10.1101/2020.03.24.20042291.

20. Lourenco, J, Pinotti, F, Thompson, C, Gupta, S. The impact of host resistance on cumulative mortality and the threshold of herd immunity for SARS-CoV-2. 2020; doi:https://doi.org/10.1101/2020.07.15.20154294.

